# Genomic ascertainment for *UBA1* variants and VEXAS syndrome: a population-based study

**DOI:** 10.1101/2022.07.27.22277962

**Authors:** David B. Beck, Dale L. Bodian, Vandan Shah, Uyenlinh L. Mirshahi, Jung Kim, Yi Ding, Natasha T. Strande, Anna Cantor, Jeremy S. Haley, Adam Cook, Wesley Hill, Peter C. Grayson, Marcela A. Ferrada, Daniel L. Kastner, David J. Carey, Douglas R. Stewart

**Affiliations:** National Human Genome Research Institute, National Institutes of Health, Bethesda, Maryland, USA; Center for Human Genetics and Genomics, New York University School of Medicine, New York, New York, USA; Division of Rheumatology, Department of Medicine, New York University School of Medicine, New York, New York, USA; Geisinger Research, North Bethesda, Maryland, USA; Department of Molecular and Functional Genomics, Geisinger Medical Center, Danville, Pennsylvania, USA; Clinical Genetics Branch, Division of Cancer Epidemiology and Genetics, National Cancer Institute, National Institutes of Health, Rockville, Maryland, USA; Department of Laboratory Medicine, Geisinger Medical Center, Danville, Pennsylvania, USA; National Institute of Arthritis and Musculoskeletal and Skin Diseases, National Institutes of Health, Bethesda, Maryland, USA

## Abstract

**Importance:** VEXAS (vacuoles, E1-ubiquitin-activating enzyme, X-linked, autoinflammatory, somatic) syndrome is a disease with rheumatologic and hematologic features caused by somatic variants in *UBA1*. Pathogenic variants are associated with a broad spectrum of clinical manifestations. Knowledge of prevalence, penetrance, and clinical characteristics of this disease have been limited by ascertainment biases based on known phenotypes. This study used a genomic ascertainment approach to overcome these limitations and better define *UBA1*-related disease.

**Objective:** Determine the prevalence of pathogenic variants in *UBA1* and associated clinical manifestations in an unselected population using a genomic ascertainment approach.

**Design, Setting and Participants:** This cohort study evaluated *UBA1* variants in exome data from 163,096 participants within the Geisinger MyCode Community Health Initiative. Clinical phenotypes were determined from Geisinger electronic health record (EHR) data up to January 1st, 2022.

**Main outcomes and measures:** Prevalence of somatic *UBA1* variation; presence of rheumatologic, hematologic, pulmonary, dermatologic, and other symptoms in individuals with somatic *UBA1* variation; structured and manual review of EHR; review of bone marrow biopsies; survival in carriers of somatic *UBA1* variation.

**Results:** In a retrospective study of 163,096 participants (mean age 52.8 years; 94% of European ancestry, 61% female), 11 individuals harbored somatic, known pathogenic *UBA1* variants, with 100% having clinical manifestations consistent with VEXAS syndrome. We found a previously unreported *UBA1* variant (c.1861A>T; p.Ser621Cys) in a symptomatic patient. Disease-causing *UBA1* variants were found in ∼1 in 14,000 unrelated individuals, and ∼1 in 4,000 men >50 years old. A disease-causing *UBA1* variant confers a ∼ 6.6 higher probability of mortality vs. age-, sex-, and BMI-matched non-carriers. The majority (7, 58%) of individuals did not meet criteria for rheumatologic and hematologic diagnoses previously associated with VEXAS syndrome, however all individuals had anemia (mean 7.8 g/dL, median 7.5g/dL), mostly macrocytic (91%) with concomitant thrombocytopenia (91%). Finally, we identified a pathogenic variant in one male prior to onset of VEXAS-related signs or symptoms and two females had disease with heterozygous variants.

**Conclusions and relevance:** This cohort study showed that the prevalence, penetrance, and expressivity of pathogenic *UBA1* variants were higher than expected. More expansive *UBA1* testing will lead to molecular diagnoses and improved treatment for patients.

## Main Text

Molecular, and specifically genetic, diagnoses are uncommon in rheumatologic diseases, limiting prognosis and treatment in many cases. The most common genetic causes of inflammation include *MEFV* and familial Mediterranean fever (FMF)^1^, *TNFRSF1A* and tumor necrosis factor receptor-associated periodic syndrome (TRAPS)^2^, and *ADA2* and deficiency of adenosine deaminase 2 (DADA2)^3,4^. These disorders are rare, typically early-onset, inherited, and clinically distinct from common rheumatologic conditions. Recently VEXAS (vacuoles, E1-ubiquitin-activating enzyme, X-linked, autoinflammatory, somatic) syndrome, arising from somatic variants in *UBA1*, was recognized as a novel disease entity^5^. Unlike previously recognized genetic causes of inflammation, *UBA1* variants are acquired and occur later in life. *UBA1* is X-linked and thus phenotypic consequences occur predominantly in men or in females with monosomy of the X chromosome^6^. Distinct germline *UBA1* variants underlie X-linked spinal muscular atrophy and occur in different regions of the protein than VEXAS syndrome^7^. Patients with VEXAS syndrome have rheumatologic, hematologic, dermatologic and pulmonary manifestations thought to be caused by inflammation and can carry a variety of clinical diagnoses including polyarteritis nodosa (PAN), relapsing polychondritis (RP), giant cell arteritis (GCA), Sweet syndrome (SS) and myelodysplastic syndrome (MDS). Prognosis in VEXAS is determined more by the molecular than clinical diagnosis^8^. Previous work on *UBA1/*VEXAS was largely phenotype-driven with ascertainment based on similarity to the original reported manifestations of VEXAS syndrome^8,9^.

Accurate diagnoses in rheumatologic diseases can be challenging due to non-specific overlapping clinical labs and disease features, need for invasive, time-sensitive biopsies, and varying natural history. We investigated the prevalence, penetrance, and expressivity of pathogenic *UBA1* variants in the DiscovEHR cohort, an unselected clinical cohort of participants in the MyCode Community Health Initiative at Geisinger^10^, a healthcare system in central and northeastern Pennsylvania. The study population consisted of the first 163,096 individuals with exome sequence and available electronic health record (EHR) data. Participation in MyCode is open to all Geisinger patients and is not based on the presence or absence of specific clinical traits. Enrollment was performed primarily during non-emergent outpatient visits in both primary care and specialty clinics. There are currently no guidelines for which individuals to test for *UBA1* mutations, and diagnostic criteria for VEXAS syndrome do not exist. Therefore, we analyzed associated clinical diagnoses, symptoms and laboratory values in an unascertained cohort to better define this disease entity.

We identified 11 individuals with known pathogenic variants in *UBA1* (NM_003334.4) that were likely somatic based on variant allele fractions (VAFs) ranging from 0.04-0.80 (median 0.23) (**Table 1, Supplemental Table 1**). Eight (73%) individuals had variants at codon 41 (c.121 A>C; p.Met41Leu, c.121A>G; p.Met41Val, c.122T>C; p.Met41Thr), two (18%) had a missense variant at codon 56 (c.167C>T; p.Ser56Phe), and one (9%) had a canonical splice site variant (c.118-2A>G). Two of the 11 variant carriers were female (18%) without evidence of aneuploidy on single nucleotide polymorphism (SNP) array or exome sequencing. Clinical *UBA1* mutation testing was not performed in any case. Upon structured data extraction and physician EHR review, four of these 11 (36%) individuals had assigned clinical diagnoses associated with VEXAS syndrome (found in 80% of the original cohort^5^) (**Table 2**). Although the minority of *UBA1* variant carriers in this cohort had a VEXAS-associated clinical diagnosis, all individuals had clinical manifestations consistent with VEXAS identified both by structured data extraction and physician EHR review (**Supplemental Table 1 and 2, Methods)** ^**5**^. All variant carriers were over 50 years old at onset of symptoms, consistent with previous reports (**Table 1)**. Separately, we identified a participant with a known pathogenic variant, *UBA1* c.118-1G>C (VAF 0.53), in the UK Biobank with overlapping clinical features consistent with VEXAS syndrome, but no classic clinical diagnosis.

**Table 1.** Demographics of *UBA1* Pathogenic Variant Carriers. Demographics for Geisinger subjects with pathogenic variants in *UBA1*. Note that some individuals were sequenced twice and both variant allele fraction (VAF) values are listed, with the higher confidence/depth of sequence highlighted in bold. VAF= Variant allele fraction. Variant annotation to cDNA used transcript ID NM_003334.4 and to protein used NP_003325.2 ID for *UBA1*. DID refers to Geisinger deidentified subject identifier.

| # | Patient ID | Sex | Deceased | Deceased Age | Age at Last Encounter | Sample Age | Disease Onset Age | HGVSc/ HGVSp | VAF |
| --- | --- | --- | --- | --- | --- | --- | --- | --- | --- |
| 1 | DID001 | Male | yes | 80's | N/A | 70's | 70's | c.121A>C; p.Met41Leu | **.085**, .075 |
| 2 | DID002 | Male | yes | 60's | N/A | 60's | 50's | c.121A>C; p.Met41Leu | 0.303 |
| 3 | DID004 | Female | yes | 70's | N/A | 70's | 60's | c.121A>G; p.Met41Val | **0.193**, 0.24 |
| 4 | DID006 | Male | no | N/A | 50's | 50's | 50's | c.167C>T; p.Ser56Phe | 0.793 |
| 5 | DID007 | Female | yes | 80's | N/A | 70's | 70's | c.118-2A>G | 0.209 |
| 6 | DID008 | Male | no | N/A | 60's | 60's | 60's | c.121A>C; p.Met41Leu | 0.742 |
| 7 | DID010 | Male | yes | 70's | N/A | 70's | 70's | c.121A>G; p.Met41Val | 0.571 |
| 8 | DID011 | Male | yes | 80's | N/A | 80's | 80's | c.167C>T; p.Ser56Phe | **0.038**, 0.16 |
| 9 | DID016 | Male | yes | 60's | N/A | 60's | 50's | c.121A>G; p.Met41Val | 0.129 |
| 10 | DID017 | Male | yes | 80's | N/A | 80's | 60's | c.122T>C; p.Met41Thr | 0.381 |
| 11 | DID018 | Male | yes | 80's | N/A | 80's | 70's | c.121A>C; p.Met41Leu | **0.52**, 0.47 |

**Table 2.** Clinical Diagnoses and Key Laboratory Values in *UBA1* Pathogenic Variant Carriers. Clinical features for Geisinger subjects with pathogenic variants in *UBA1*. Hematologic involvement defined as anemia and/or thrombocytopenia. Skin and pulmonary diagnoses listed when possible, and with details for ICD coding (Supplemental Table 1) and clinical diagnoses (Supplemental Table 2). Classic % are results originally reported for *UBA1*/VEXAS based on diagnoses in the original report.^5^ Laboratory values references ranges: Hemoglobin (Hgb)= 12-17 g/dL; Platelets (Plt)= 150-400 × 10^9^/L; Mean Corpuscular Volume (MCV)= 80-95 fL; *= median; ^=mean. DID refers to Geisinger subjects.

|  |  | Diagnoses |  |  |  |  | Clinical Characteristics |  |  |  | Laboratory Values |  |  |  |
| --- | --- | --- | --- | --- | --- | --- | --- | --- | --- | --- | --- | --- | --- | --- |
| # | Patient ID | Rheumatologic diagnoses | Relapsing Polychondritis | Sweet Syndrome | Polyarteritis Nodosa | Myelodysplastic Syndrome | Arthritis | Hematologic Involvement | Skin involvement | Pulmonary involvement | Hgb (nadir) | MCV (peak) | Plt (nadir) | Bone Marrow Biopsy Results |
| 1 | DID001 | No | No | No | No | No | Yes | Yes | No | Yes | 7.4 | 93.1 | 91 | Cytoplasmic vacuoles (myeloid) and megaloblastic changes |
| 2 | DID002 | No | No | No | No | No | No | Yes | No | Yes | 6 | 119 | 28 | Cytoplasmic vacuoles (myeloid) and megaloblastic changes |
| 3 | DID004 | Psoriasis, Polymyalgia Rheumatica | No | No | No | No | Yes | Yes (CML, MGUS) | No | No | 7.5 | 126 | 18 | Absent |
| 4 | DID006 | No | No | No | No | Yes | Yes | Yes (Neutropenia, Myelo-fibrosis) | No | Yes | 5.5 | 100.9 | 8 | Cytoplasmic vacuoles (myeloid and erythroid) and megaloblastic changes |
| 5 | DID007 | No | No | No | No | No | Yes | Yes | No | Yes | 9 | 97.8 | 178 | N/A |
| 6 | DID008 | Dermatomyositis | No | No | No | Yes | Yes | Yes | Yes | No | 7.5 | 116 | 55 | Cytoplasmic vacuoles (myeloid) and megaloblastic changes |
| 7 | DID010 | Lofgren's Syndrome | No | No | No | Yes | Yes | Yes | Yes (Erythema Nodosum) | No | 7.3 | 109.3 | 102 | Cytoplasmic vacuoles (myeloid) |
| 8 | DID011 | No | No | No | No | No | No | Yes | No | Yes | 7.9 | 97.3 | 83 | N/A |
| 9 | DID016 | No | No | No | No | No | Yes | Yes | Yes | Yes | 10.2 | 115.5 | 82 | N/A |
| 10 | DID017 | Giant Cell Arteritis | No | No | No | No | No | Yes | Yes | Yes | 7.8 | 126.4 | 118 | N/A |
| 11 | DID018 | No | No | No | No | No | Yes | Yes | Yes | No | 10 | 115.8 | 125 | N/A |
|  | **Total %** | 36.4 | 0.0 | 0.0 | 0.0 | 27.3 | 63.6 | 100.0 | 45.5 | 63.6 | **\*7.5** | **\*115.5** | **\*83** |  |
|  | **Classic %** | 76 | 60 | 32 | 12 | 34 | NA | 96 | 88 | 72 | **^7.8** | **^111.1** | **^80.9** |  |

Seven (64%) had arthritis and four (36%) were diagnosed with rheumatologic diseases including psoriasis, polymyalgia rheumatica, dermatomyositis, and Lofgren syndrome (**Table 2**). Only four (36%) individuals had features on chart review consistent with reported VEXAS flares (fevers, elevated acute phase reactants, steroid dependency) (**Supplemental Table 1 and 2**). All 11 (100%) had anemia, ten (91%) with macrocytosis, five (46%) required transfusions, ten (91%) with thrombocytopenia, five (46%) with pancytopenia with anemia, thrombocytopenia, and lymphopenia, three (27%) with MDS and one (9%) with chronic myelogenous leukemia (CML) (**Table 2 and Supplemental Table 2**). Five (46%) had skin involvement and seven (64%) had pulmonary disease. A progressive increase in mean corpuscular volume (MCV) was noted, with macrocytosis only developing after first symptoms onset (data not shown). Bone marrow biopsies were performed in six patients, with hypercellular marrow detected in all cases. Cytoplasmic vacuoles in myeloid and/or erythroid precursors were observed in five (83%) patients, while megaloblastic changes in erythroid elements were seen in four patients (67%). (**Table 2, Supplemental Table 2)**. Moreover, 80% (4/5) of those tested showed elevated ESR (**Supplemental Table 2**).

Nine of the 11 variant carriers are deceased (82%), and survival analysis showed a significant relationship between harboring a known pathogenic *UBA1* variant and mortality (concordance 0.8, p=0.02). Unrelated *UBA1* pathogenic variant carriers were more likely to die than age- and sex-matched unrelated noncarriers (HR=6.6 [95% CI 1.8-24.4]) (**Figure 1A, Supplemental Table 3**). There was no statistically significant relationship between VEXAS disease duration and mortality in *UBA1* carriers, likely due to sample size (concordance 0.6 +/- 0.1 standard deviation, p=0.7. **Figure 1B**).

**Figure 1.**
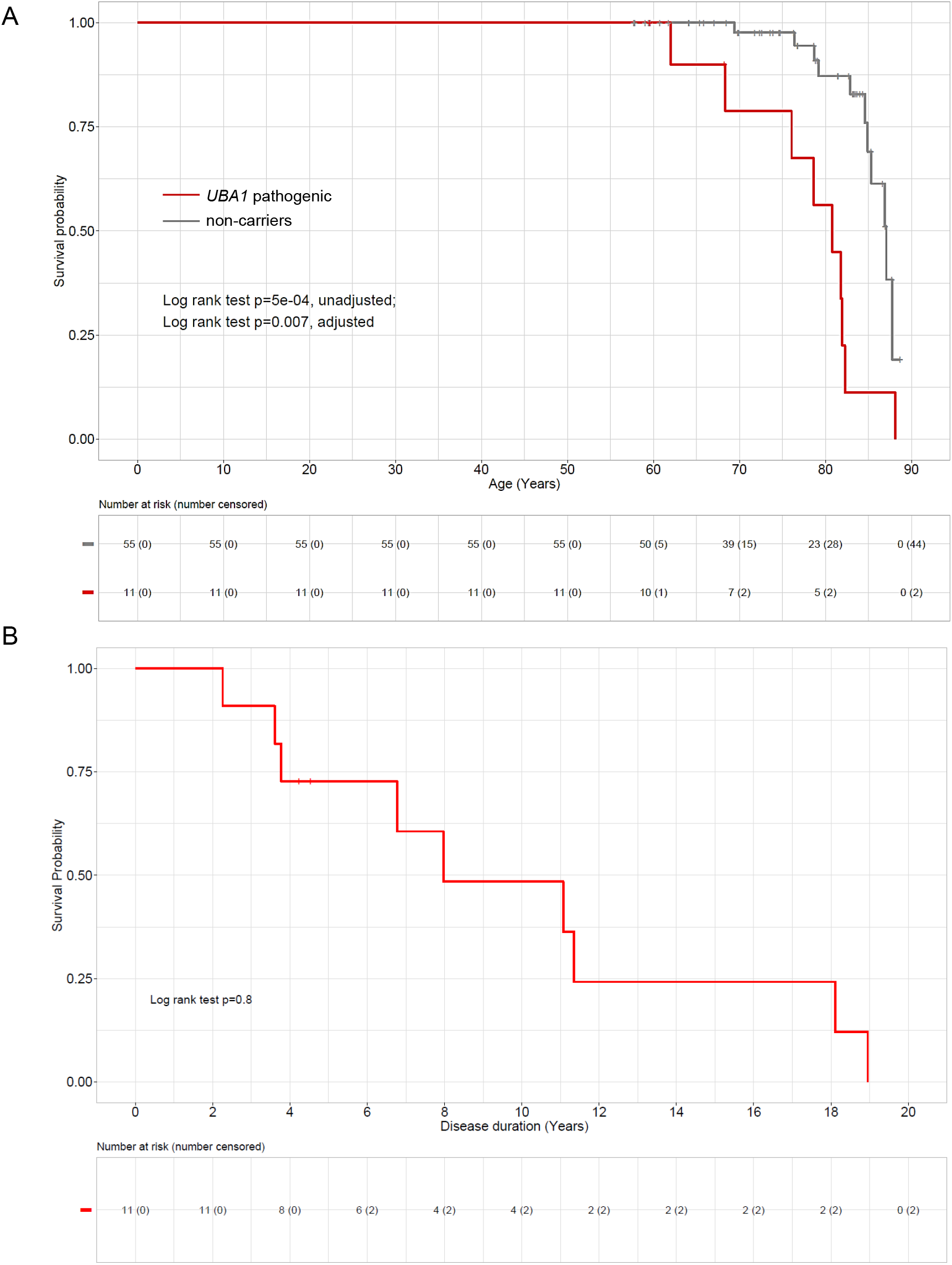
Survival analyses of individuals with *UBA1* pathogenic variants. **A**. Cumulative survival of pathogenic *UBA1* carriers relative to noncarriers. Kaplan-Meier survival curves for unrelated carriers of known pathogenic *UBA1* variants (red) and unrelated noncarriers (gray) matched by ancestry, sex, and age of the earliest encounter with a Geisinger healthcare provider at a random 1:5 carrier:noncarrier ratio. Censored subjects were marked with a | on the curves. The risk table indicates the number of individuals in each group at the start of the study and the number remained at each decade. The number of censored subjects at each decade are in parentheses. Log-rank test p values showed statistical significance between the curves with and without adjustment for sex, earliest age of encounter, and body mass index. **B**. Survival of *UBA1* carriers following VEXAS presentation. Kaplan-Meier survival curves for unrelated carriers of pathogenic *UBA1* variants following VEXAS disease onset.

We also queried for novel disease-causing *UBA1* variants in males and identified three rare, likely somatic, missense or splicing variants in three males greater than 50 years old (**Supplemental Table 4**). Although none of these males had documented clinical diagnoses typically associated with VEXAS syndrome, one individual (DID005, c.1861A>T; p.Ser621Cys, VAF 0.2. Patient identifiers are not known to anyone outside of the research group) had clinical characteristics consistent with VEXAS syndrome with a clinical diagnosis of granulomatosis with polyangiitis (GPA). We therefore consider this variant to be highly suspicious for causing VEXAS syndrome.

We identified 10 males and 2 females with either pathogenic (11) or highly suspicious disease-causing variants (1), giving an estimated variant prevalence of 1/4,000 for males and 1/26,000 for females, or 1/8,000 combined for individuals age >50 years. The estimated prevalence is ∼1/14,000 for the entire cohort. Our findings demonstrate that *UBA1* pathogenic variants are more common than expected, can precede clinical onset, and appear to be highly penetrant. *UBA1* pathogenic variants may be more common than the reported prevalence of clinical diagnoses comprising VEXAS syndrome including most vasculitides (GPA ∼1/18,000, PAN ∼1/33,000)^11,12^ and has a prevalence similar to Behçet’s Disease (∼1/10,000) and MDS (∼1/14,000)^13,14^. Although these clinical diagnoses are likely under-recorded because of diagnostic and physician-coding challenges and cohort specific approaches, our reported *UBA1* variant prevalence is also likely underestimated due to detection limits of research-grade exome data, use of peripheral blood over bone marrow samples, and the possibility of novel and as-yet-unrecognized pathogenic variants. Our work is limited to care provided within a single center, and cannot exclude clinical findings, treatment or testing provided by an external provider.

Many of the classic features and diagnoses previously recognized as VEXAS syndrome in the literature (RP, PAN, Sweet syndrome) were not identified in the Geisinger cohort, except for MDS. Thus, given our current understanding of VEXAS, many of these patients may not have been identified using phenotype-based ascertainment. Our findings expand the clinical spectrum of VEXAS to include individuals over the age of 50 years without specific clinical rheumatologic diagnoses, but experiencing a combination of rheumatologic, hematologic, dermatologic, or pulmonary symptoms along with anemia, thrombocytopenia, and elevated inflammatory markers. We also demonstrate for the first time that the variant can precede onset of clinical symptoms. Further highlighting the strength of an unbiased genomic ascertainment approach to identify patients with VEXAS from within the general population, we observed a different proportion of VEXAS-defining mutations compared to two recent phenotypically defined cohort studies in VEXAS^8,9^, and we identified an additional likely disease-causing variant. Finally, we also observed a greater proportion of euploid females with pathogenic variants in *UBA1* than previously reported.^15^

Despite the large number of cases reported to date, *UBA1* is still not routinely offered on standard workup for myeloid neoplasms or immune dysregulation diagnostic panels ^8,9^. Defining the clinical spectrum of *UBA1*, using biobanks, is a necessary step to better define disease criteria and testing recommendations. Identification of *UBA1* pathogenic variants defines VEXAS syndrome, and can alter treatment, prognosis and screening plans ^16,17^, therefore we propose broad *UBA1* testing in patients with features indicative of VEXAS, including those with non-specific differential diagnoses, such as macrocytic anemia of uncertain etiology in association with elevated inflammatory markers.

## Data Availability

All data produced in the present study are available upon reasonable request to the authors

## Supplemental Methods

### Sequencing/Alignment

Exome sequencing and variant calling were performed in collaboration with Regeneron Genetics Center as described previously^18^. NimbleGen probes (SeqCap VCRome) and xGEN probes from Integrated DNA Technologies (IDT) and were used for target sequence capture. Sequencing was performed by paired end 75bp reads on an Illumina NovaSeq or HiSeq at coverage >20x at >80% of the targeted bases. Alignments and variant calling were based on GRCh38 human genome reference sequence. Variants were called with the WeCall variant caller version 1.1.2. (https://github.com/Genomicsplc/wecall).

Pathogenic (P) and likely pathogenic (LP) variants for *UBA1*, including those listed both for somatic and germline diseases, were extracted from ClinVar (https://www.ncbi.nlm.nih.gov/clinvar/). No pathogenic variants for XL-SMA were identified within the cohort. All genotype calls of P and LP variants supported by >=3 alt reads with genotype quality >=30 were selected for analysis. Two of the patients have low VAF (<0.10), which can be associated with lower confidence genotype calls. However, both were sequenced twice, on two different exome platforms, and detected in both sequencing results, and both patients had symptoms consistent with VEXAS. Low VAF mutation was identified in DID001 using mutect2, and manual analysis, although this variant caller was not used for the entire cohort.

### Electronic Health Record (EHR) and Pathology Review

EHR review was performed by structured review and manual review of all available medical records within the Geisinger health care system. Longitudinal clinical data spanning 4-25 years (median 12 years) was reviewed both by Geisinger phenomic analytics and clinical data core and practicing internal medicine physicians (**Supplemental Table 1**). Structured EHR review included review of all assigned ICD10 visit diagnoses, all prescribed medications, and any procedures performed. Consultations with rheumatology, dermatology, pulmonology, and hematology were recorded. All data from the following laboratory tests were collected and analyzed: complete blood count, erythrocyte sedimentation rate, and C-reactive protein. All structured data related to VEXAS syndrome was included in the manuscript, after manual review of complete raw structured data (**Supplemental Table 1**). Manual chart review was performed by an internal medicine physician using a standard case form report and clinical data was detailed in de-identified patient-level descriptive summaries. Centralized review of peripheral blood and bone marrow aspirate smears, histologic sections, immunohistochemical and cytogenetic studies performed at the time of diagnosis were performed by a pathologist.

Given the lack of VEXAS syndrome ICD10 code or diagnostic criteria, a *UBA1* carrier was considered to have VEXAS syndrome if there were concomitant clinical features in two different organ systems known to be associated with VEXAS syndrome (rheumatologic, hematologic, dermatology, pulmonary) along with supportive laboratory abnormalities (anemia).

Disease onset date refers to first symptoms related to VEXAS syndrome mentioned in the chart. Distinct day counts, refers to number of days individuals were seen at Geisinger. Given that patients were examined at different times, by different physicians, it is possible that clinical diagnoses were present but not assessed or accurately reported. Institutional review board (IRB) of Geisinger Medical Center gave ethical approval for this work.

### Survival Analysis

Kaplan-Meier survival analysis was used to estimate the probability of mortality for *UBA1* carriers relative to noncarriers. Samples of participants up to age 90 years were included. Data were left-censored to the age of the earliest encounter with a Geisinger provider plus 3 months, and right-censored to the age of the last encounter with a Geisinger provider. Carriers and noncarriers were unrelated up to third-degree relationship and matched by European ancestry, sex, and the earliest age of encounter + 1 year. Cox proportional hazard regression was used to compute hazard ratios and 95% confidence intervals (95% CI) with or without adjustment for earliest age of encounter, sex, and body mass index. Regression was performed on all matched individuals and a random 1:5 carrier: noncarrier ratio. Concordance is defined as 80% of data fit the survivor function equation.

### Variant of Uncertain Significance Query

Variants of uncertain significance were defined as missense or consensus splice site variants detected in males with age of blood sample collection >50 years, not found in any annotated relative and in <=4 DiscovEHR participants with complete sample collection data. They were filtered for number of reads supporting the alternate allele >=3, Depth (DP) >=20, variant allele fraction (VAF) >=0.2, genotype quality (GQ)>=50, and allele frequency in gnomAD <1e-05. Variants with VAF>0.8 in all males and VAF>= 0.4 and VAF <0.6 in all females were excluded.

## Acknowledgements

This research has been conducted using the UK Biobank Resource under Application Number 54389. This research was supported by Intramural Research Program of the Division of Cancer Epidemiology and Genetics of the National Cancer Institute and of the National Institutes of Health. D.B.B. was supported by the Relapsing Polychondritis Foundation and funding from the NIH (R00AR078205). We are also thankful to Matt Oetjens for mosaicism analysis in genotyping data. Support for MyCode enrollment and exome sequencing were provided by the Regeneron Genetics Center.

## Author contributions

D.B.B., D.J.C., D.R.S. designed the study. D.B.B., D.L.B., V.S. U.L.M. J.K. Y.D., N.S., A.C., J.S.H., A.C., W.H., gathered data. D.B.B., D.L.B., V.S. U.L.M. J.K. Y.D., N.S., A.Ca., J.S.H., A.C., W.H, P.C.G, M.A.F., D.L.K, D.J.C., D.R.S. analyzed data.

## Figure Legends

**Supplemental Table 1. Electronic Health Queries for individuals with *UBA1* pathogenic variants**. Distinct day count refers to number of days seen within Geisinger healthcare system and duration of care refers to the number of years visits covered.

**Supplemental Table 2. Structured and manual review of EHR for individuals with *UBA1* pathogenic variants**. Detailed data about clinical symptoms and additional laboratory values in addition to Table 2. Erythrocyte Sedimentation Rate (ESR) value upper level of normal= 15mm/hr.

**Supplemental Table 3. Univariate and multivariate Cox proportional regression hazard ratios for mortality of individuals with *UBA1* pathogenic relative to noncarriers**. Pathogenic *UBA1* variant carriers and noncarriers were matched by European ancestry, sex, and age of the earliest encounter with a Geisinger healthcare provider. All individuals were unrelated to each other in each group up to third-degree. Hazard ratios (95% CI), regression concordance (Standard deviation), p values, and the number of unrelated carriers in univariate (*) and multivariate (adjust §) regression analysis vs. unrelated noncarriers was performed on all matched individuals or on a random 1:5 carrier: noncarrier ratio. Multivariate regression adjusted for age of the earliest encounter, sex, and body mass index.

**Supplemental Table 4. Variants of Uncertain Significance in *UBA1* and associated phenotypes based on structured and manual review of EHR**. Demographics for Geisinger subjects with variants of uncertain significance in *UBA1*. VAF= Variant allele fraction. Transcript (NM_003334.4) and protein (NP_003325.2) ID for UBA1 were used for HGVSc/HGVSp.

## Notes

### Competing Interest Statement

The authors have declared no competing interest.

### Author Declarations

Institutional review board (IRB) of Geisinger Medical Center gave ethical approval for this work.

